# Cell-free urine- and plasma DNA mutational analysis predicts neoadjuvant chemotherapy response and outcome in patients with muscle invasive bladder cancer

**DOI:** 10.1101/2022.10.24.22281440

**Authors:** Emil Christensen, Iver Nordentoft, Sara K. Elbæk, Karin Birkenkamp-Demtröder, Ann Taber, Tine G. Andreasen, Trine Strandgaard, Michael Knudsen, Philippe Lamy, Mads Agerbæk, Jørgen B. Jensen, Lars Dyrskjøt

## Abstract

**Purpose:** Investigate and compare the use of plasma- and urine DNA mutation analysis for predicting neoadjuvant chemotherapy (NAC) response and long-term oncological outcome in patients with muscle invasive bladder cancer.

**Experimental Design:** Whole exome sequencing of tumor and germline DNA was performed for 92 patients treated with NAC followed by radical cystectomy (RC). A custom NGS panel capturing approx. 50 mutations per patient was designed and utilized to track tumor-derived DNA (tdDNA) in liquid biopsies. A total of 447 plasma samples, 281 urine supernatants and 123 urine pellets collected before, during and after treatment were analyzed. Patients were enrolled from 2013-2019 with a median follow-up time of 41.3 months after RC.

**Results:** We identified tdDNA before initiation of NAC in 89% of urine supernatants, 85% of urine pellets and 43% of plasma samples. tdDNA levels were higher in urine supernatants and urine pellets compared to plasma samples (*p*<0.001). In plasma, detection of tdDNA before NAC was associated with a lower NAC response rate (p<0.001). Detection of tdDNA after NAC was associated with lower response rates in plasma, urine supernatant and urine pellet (p<0.001, p=0.03, p=0.002). tdDNA dynamics during NAC was predictive of NAC response and outcome in urine supernatant and plasma (*p*=0.006, *p*=0.002). A combined measure from plasma and urine supernatant tdDNA dynamics stratified patients by outcome (*p*=0.003).

**Conclusions:** Analysis of tdDNA in plasma and urine samples both separately and combined has potential to predict treatment response and outcome.

## Introduction

Localized muscle-invasive bladder cancer (MIBC) is a common malignancy with approx. 570,000 cases diagnosed globally in 2020(1). The standard treatment regimen is neoadjuvant chemotherapy (NAC) followed by radical cystectomy (RC), however, approx. 50% of patients experience disease recurrence following surgery(2). The current NAC response evaluation consists of a pathological assessment of the surgically removed bladder and adjacent lymph nodes and thereby inherently lacks the potential to identify patients where a change in treatment regimen might be favorable. Recent studies have identified improved outcomes for patients responding to NAC, in particular if pathological complete response (pCR, pT0N0) is achieved(3). However, meta-analyses have observed pCR in only approx. 25% of patients and pathological downstaging (=<pTisT0TaN0) in approx. 50% and considerable overtreatment might therefore be taking place(4,5). In addition, a large meta analysis identified a 5% improved survival rate for patients treated with NAC compared to non-treated patients(6). Studies employing a tumor-centric approach for prediction of NAC response have identified genomic alterations in DNA damage repair pathways and genomic instability to be associated with an increased likelihood of response(7–10). Gene expression subtypes have also been associated with NAC response, however, conflicting results have been reported (10–13). No molecular markers have yet entered into clinical practice, although a clinical trial aiming to demonstrate an association between NAC response and mutations in DNA damage repair pathways is ongoing (NCT03609216).

Circulating tumor DNA (ctDNA) has emerged as a powerful biomarker reflecting tumor invasiveness and patient outcome in multiple cancers, including MIBC(14–17). We recently demonstrated that ctDNA dynamics in plasma samples during treatment with NAC reflect treatment response, indicating that ctDNA measurements during treatment could provide a measure of response before RC is carried out(18). A study by Chauhan *et al*. investigated urinary tumor derived DNA (tdDNA) from samples collected before RC and identified higher levels among patients without NAC response compared to those with response(19). Patel *et al*. have similarly demonstrated persistence of urine based tdDNA during NAC to be associated with recurrence after RC(20). However, previous studies focusing on NAC response and urine samples have been conducted in small cohorts. Here we present a larger study on 92 patients with MIBC focusing on analysis of tdDNA in paired plasma and urine samples to investigate the potential for NAC response prediction.

## Materials and Methods

Additional information is available in Supplementary Data.

### Patient cohort

A total of 92 patients treated with NAC before RC were prospectively enrolled between 2014 and 2019 at Aarhus University Hospital, Denmark. Treatment and surveillance was done in accordance with Danish national guidelines, as previously described(18). Whole exome sequencing (WES) and plasma data for 56/92 patients was generated previously(18). Pathological downstaging, as evaluated from the RC specimen, was defined as pT0/Ta/TisN0. Only patients with plasma and/or urine supernatants and/or urine pellets were included in the study. Detailed follow-up data was available for all patients (Supplementary Table 1). Recurrence data was obtained from computed tomography scans or pathology reports and survival data was obtained from the nationwide civil registry. All patients provided informed written consent, and the study was approved by The National Committee on Health Research Ethics (#1302183). Study data were collected and managed using REDCap hosted at Aarhus University(21,22).

### Clinical samples

Tissue samples for WES were obtained from transurethral resection of the bladder (TURB) at the time of diagnosis (n= 90) or from RC specimens (n=2). In addition, 32 tissue samples from RC specimens, with paired TURB tissue available, were included for mutation tracking over time. DNA was extracted from sections of Tissue-Tek^?^ O.C.T Compound embedded tissue or punches of formalin-fixed paraffin embedded tissue (FFPE) using Puregene DNA purification kit (Gentra Systems), QIAamp DNA FFPE tissue kit or Allprep DNA/RNA Kit (QIAGEN). Leukocyte DNA was extracted from the buffy coat from all patients using the QIAsymphony DSP DNA midi kit (QIAGEN). Urine and plasma sample processing, storage and DNA extraction was performed as previously described(18,23,24). A median of 4 mL plasma (range: 2.5-4 mL) and 4 mL urine supernatant (range: 2.9-5 mL) were used for extraction of cell-free DNA (cfDNA) yielding a median of 9.0 ng/mL for plasma samples and 1.7 ng/mL for urine supernatants.

### Liquid biopsy sequencing and tumor DNA detection

The patient cohort was split in two for custom panel design and a panel was designed based on WES data for each subcohort. Panel 1 covered a total of 2474 unique genomic positions (50 patients), resulting in 50 positions of interest per patient. Panel 2 covered a total of 2087 unique genomic positions (42 patients), resulting in 43-50 positions of interest per patient. DNA extracted from liquid biopsies was subjected to Twist Bioscience Mechanical Fragmentation Library preparation.

However, the adapters were replaced with xGen™ UDI-UMI adapters (Integrated DNA Technologies) in order to incorporate unique molecular identifiers (UMIs) for reducing the error rate. DNA extracted from urine pellets was fragmented prior to library preparation to a fragment size of approx. 350 bp using the Twist Library Preparation Enzymatic Fragmentation (EF) Kit 1.0 (Twist Bioscience). A median DNA input of 39.5 ng (range: 5.7-258 ng), 50 ng (range: 4-50 ng) and 7.6 ng (range: 0.2-110 ng) was used for library preparation for plasma samples, urine pellets and urine supernatants, respectively. The enrichment process was carried out using Twist Bioscience Target Enrichment Protocol and the described custom panels. The libraries were paired end sequenced (2×150 bp) on the illumina NovaSeq 6000 sequencer. UMI consensus base calls were performed using the fgbio tool package(25). Samples were sequenced to median target coverages of 3346X, 1560X and 2588X for plasma, urine pellet and urine supernatant, respectively, after UMI consensus collapsing. The overlapping parts of read pairs were only counted once. In this work we apply the following nomenclature for simplicity: tumor derived DNA (tdDNA) and this covers circulating tumor DNA (ctDNA) for mutated plasma DNA; urinary tumor DNA (utDNA) for mutated urine supernatant DNA; tumor derived DNA (tdDNA) for mutated DNA from urine pellets.

### Statistical analyses

Statistical significance was assessed by performing a Wilcoxon rank sum test for continuous non-paired variables, a Wilcoxon signed rank test for continuous paired variables and a Fisher’s exact test for categorical variables. Sample level assessment of tdDNA status was performed using Fisher’s method. Survival analyses were carried out in R using packages survminer and survival (https://cran.r-project.org).

### Data availability

The raw sequencing data generated in this study are not publicly available as this compromise patient consent and ethics regulations in Denmark. Processed non-sensitive data are available upon reasonable request from the corresponding author.

## Results

### Patients and methodology

In total, 92 patients treated with NAC for MIBC were included in the study. Ninety-one patients underwent RC after NAC and 61% showed pathological downstaging (53% complete response, pT0N0). Median follow-up after RC was 41.3 months. Tumor and germline DNA was subjected to WES for identification of somatic variants with a median target coverage of 114X for tumor and 73X for germline samples. The study builds on previously published tissue-based WES and plasma sample based data for 56 patients(18). Here, we extended the cohort with an additional 36 patients with plasma based analyses (n = 159) and included urine supernatant (n = 281) and urine pellet (n = 123) analyses for all patients. For the extended cohort and new samples, we employed a tumor-informed patient specific strategy for detection of tdDNA in liquid biopsies based on selection of approx. 50 mutations per patient from WES data for design of a custom NGS panel (Fig. 1a). Plasma and urine-based samples were subsequently subjected to targeted sequencing. Analysis of tdDNA was performed on a mutation-wise level based on error-models of all relevant genomic positions and an integrated sample-level assessment for tdDNA positivity (Fig. 1b). The limit of detection was determined to be approx. 0.05%, 0.10% and 0.09% for plasma, urine pellet and urine supernatant, respectively (Supplementary Data and Supplementary Fig. 1).

**Fig 1.**
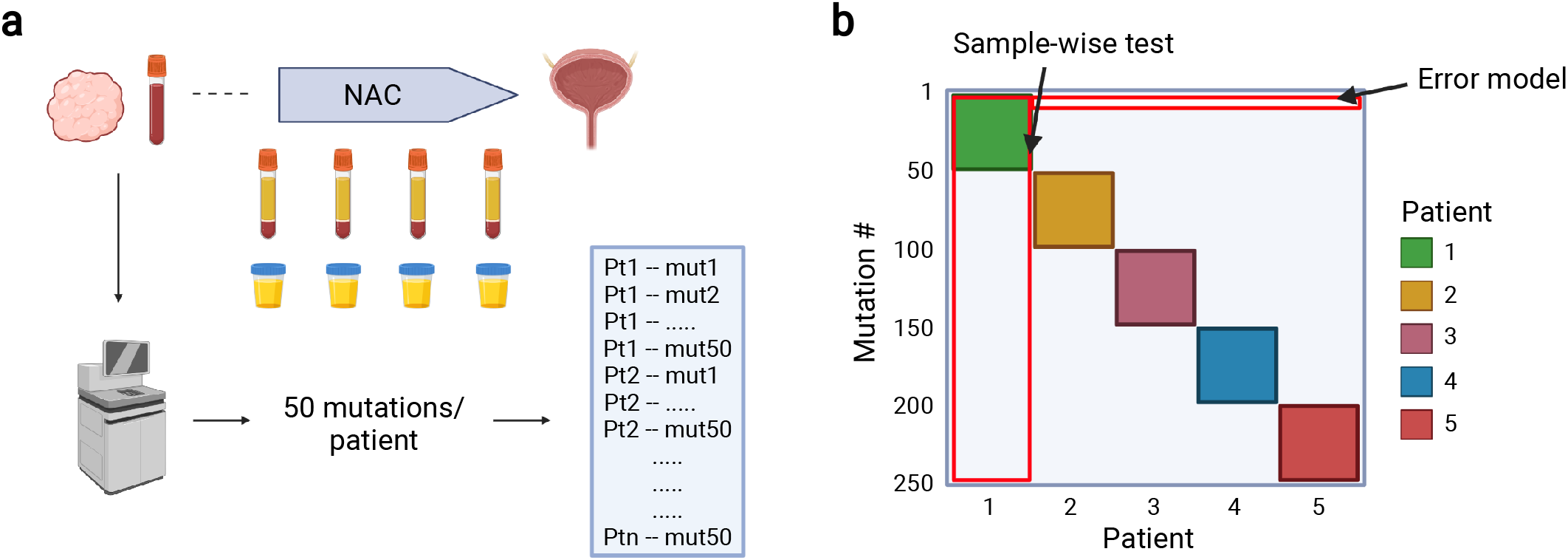
Study and methodology overview. a) The study includes patients treated for muscle invasive bladder cancer (MIBC). Upon diagnosis, a tumor sample obtained from transurethral resection of the bladder and a germline sample were subjected to whole exome sequencing (WES). Approx. 50 somatic variants were selected per patient for design of two custom NGS panels based on 50 and 42 patients. Plasma and urine samples were collected before, during and after treatment with neoadjuvant chemotherapy. b) Illustration of the custom panel NGS data, exemplified using only five patients. For every single mutation an error model was constructed based on data from the samples not associated with that given mutation. This was used to test for significance in the sample associated with that given mutation. Furthermore, a sample-wise test was performed for every sample by applying Fisher’s method to p-values obtained for all mutations associated with that sample. A random selection of 50 other mutations from the panel not associated with the given sample was similarly assessed using Fisher’s method. This was repeated 10,000 times and only samples with a score higher than all random selections were considered tdDNA positive. Created with biorender.com.

### Tumor DNA detection across sample types

The tdDNA detection frequency varied between sample types and was higher in urine supernatants and pellets compared to plasma samples (Fig. 2a). For tdDNA tracking purposes (tdDNA dynamics), detection of tdDNA before initiation of NAC is necessary and here we similarly observed tdDNA to be present more frequently in urine supernatants and pellets compared to plasma samples (Fig. 2b). In line with this observation, we found the level of tdDNA to be higher in urine supernatants and pellets compared to plasma samples both when considering all samples and only samples before NAC (Fig. 2c-d).

**Fig 2.**
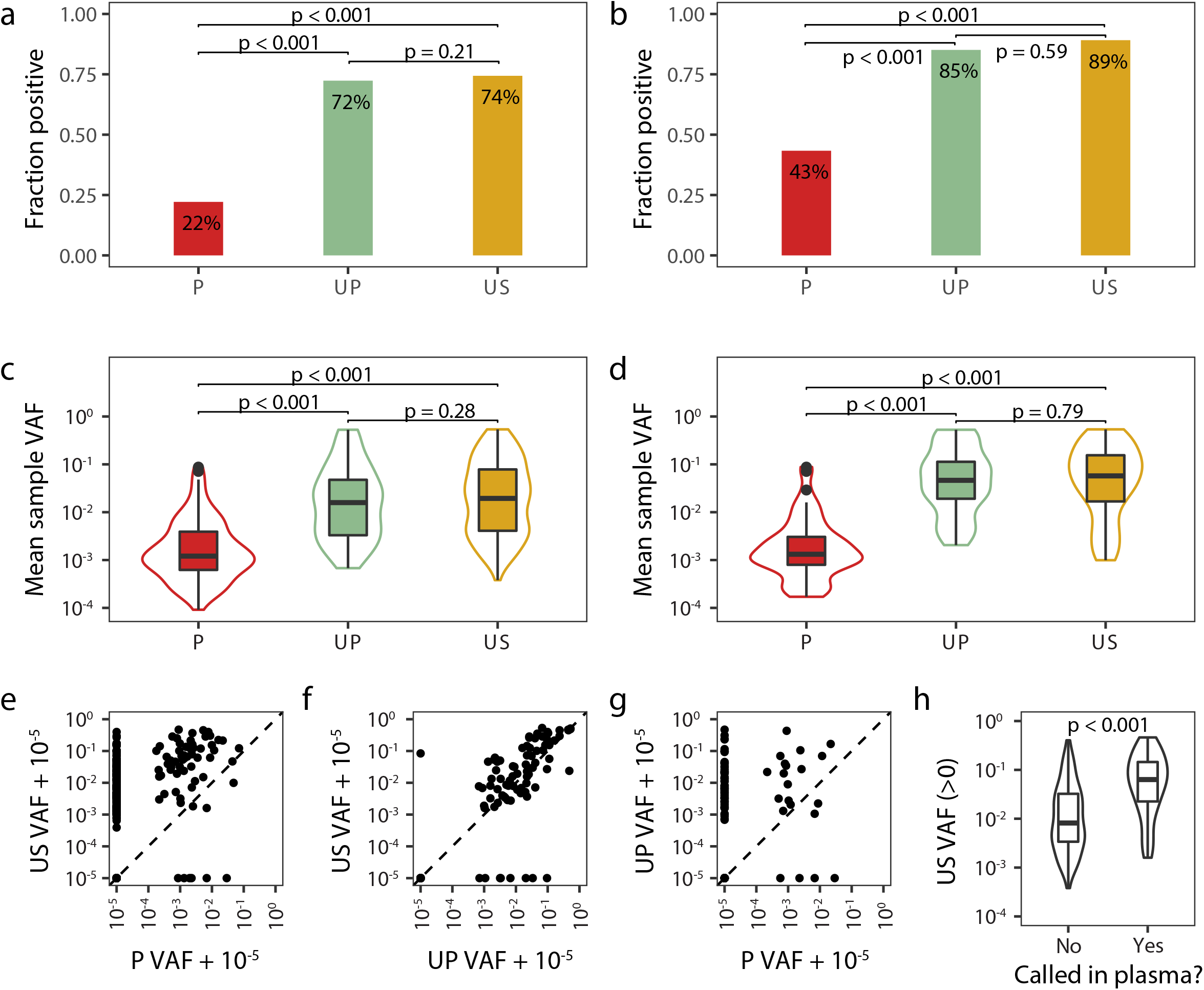
Measurements of tumor-derived DNA across liquid biopsy sample types. a-b) The fraction of samples positive for tumor-derived DNA (tdDNA) by sample type. a) All samples and b) samples collected before neoadjuvant chemotherapy (NAC). c-d) Mean sample VAF levels for samples with detectable tdDNA split by sample type. c) All samples and d) samples collected before NAC. P values were calculated using a wilcoxon rank sum test. e-g) Mean sample VAF levels for comparison of all sample types. Only samples from clinical visits with at least 2 sample types available were included. h) Mean sample VAF level for urine supernatants split by tdDNA call status in plasma samples from the same clinical visit. Only samples with detectable tdDNA in urine supernatants were considered. P values were calculated using a wilcoxon rank sum test. P = Plasma, UP = Urine pellet, US = Urine supernatant.

Urine supernatants and plasma demonstrated a weak correlation and a sample concordance in tdDNA status of 46.0%, with urine supernatants more often containing tdDNA compared to plasma (rho = 0.41, Fig. 2e). tdDNA levels in urine supernatants and urine pellets were highly correlated and 91.7% of samples were concordant in terms of tdDNA status (rho = 0.78, Fig. 2f).

Urine pellet tdDNA status also showed a weak correlation to plasma, as expected (43.1% concordance, rho = 0.15, Fig. 2g). Interestingly, for visits with tdDNA presence in plasma, the associated urine supernatants displayed significantly higher levels of tdDNA (Fig. 2h), which could be an indication of renal clearance of tdDNA from the plasma or simply a reflection of a large invasive tumor. However, no other comparisons with adjustment for tdDNA status were significant (Supplementary Fig. 2).

We observed highly variable amounts of cfDNA across sample types. However, the available amount of cfDNA used as library input was not associated with the tdDNA fraction or the number of variants detected per sample, however, the amount of extracted cfDNA was associated with the tdDNA call status of plasma samples (Supplementary Fig. 3 & 4).

We analyzed urine dipstick data collected at the same time as urine supernatants in order to investigate urine parameters impacting DNA mutation calling. Based on 72 cases of paired urine supernatant and urine dipstick data, we identified the levels of leukocytes, nitrite, protein and erythrocytes to be associated with the level of cfDNA (Supplementary Fig. 5). However, only the level of erythrocytes was associated with the tdDNA positive call status of samples and the level of tdDNA (Supplementary Fig. 6 & 7), indicating that DNA mutation calling was not affected with wild type DNA contamination. Furthermore, it indicated that patients with blood in the urine may have a higher tumor burden.

### tdDNA measurements compared to tumor burden and patient outcome

We observed ctDNA plasma sample positivity prior to initiation of NAC in 25% and 46% of visits with concurrent stage T1 and T2-T4 tumors, respectively (Fig. 3a). In the setting after NAC, only samples drawn from patients with muscle-invasive tumors at the time of RC were positive for plasma ctDNA (Fig. 3b). In urine supernatants, utDNA sample positivity rates before NAC were 100% and 90% for stage T1 and T2-4 tumors, respectively, (Fig. 3a), and varied across tumor stages after NAC (Fig. 3b). In urine pellets, we observed positive fractions similar to urine supernatants both before and after NAC (Fig. 3b).

**Fig 3.**
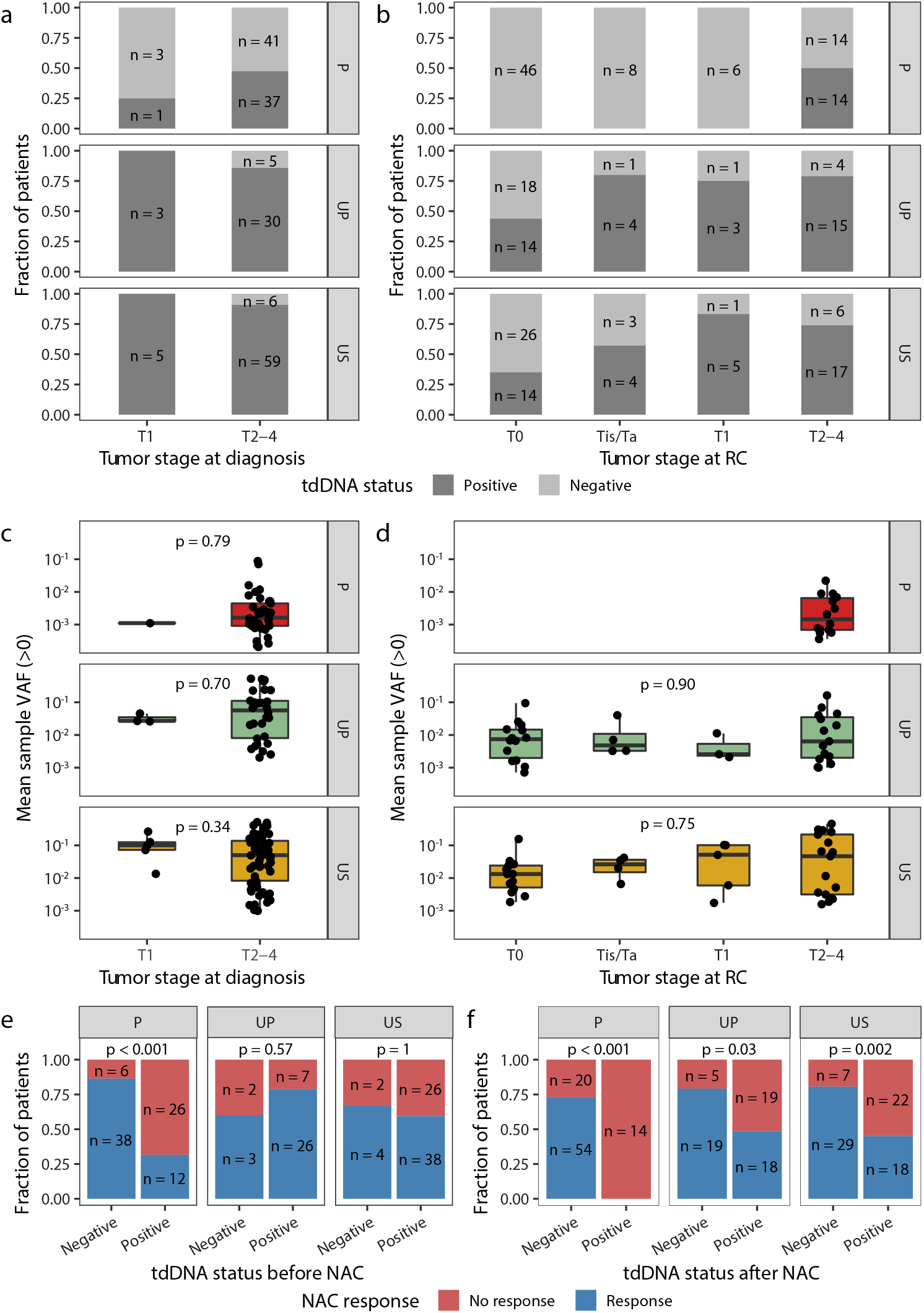
Tumor-derived DNA status before and after neoadjuvant chemotherapy compared to tumor stage and outcome. a-b) Fraction of patients positive for tumor-derived (tdDNA) split by sample type and tumor stage. a) Tumor stage evaluation based on transurethral resection of the bladder (TURB) specimen and tdDNA status based on samples collected before NAC. b) Tumor stage evaluation based on radical cystectomy (RC) specimen and tdDNA status based on samples collected after neoadjuvant chemotherapy (NAC) and before RC. c-d) Mean sample VAF levels for all patients with detectable tdDNA split by sample type and tumor stage. c) Tumor stage evaluation based on TURB specimen and tdDNA level based on samples collected before NAC. P values were calculated using a Wilcoxon rank-sum test. d) Tumor stage evaluation based on RC specimen and tdDNA level based on samples collected after NAC and before RC. P values were calculated using a Kruskal-Wallis test. e-f) tdDNA status compared to response to treatment with NAC for all analyzed sample types. e) tdDNA status determined before NAC. f) tdDNA status determined after NAC.

In urine-based samples, the levels of utDNA and tdDNA were equal across tumor stages. This could be due to urine-based tumor DNA primarily reflecting tumor exposure in the bladder lumen and less so the invasiveness of the tumor (Fig. 3c-d). In line with this, tumor DNA was always detected in both urine supernatants and pellets, when only considering samples collected before TURB (Supplementary Fig. 8). In addition, we observed an association between NAC response and plasma ctDNA status both before and after NAC, and only after NAC for urine supernatants and urine pellets (Fig. 3e-f). Only plasma ctDNA status before and after NAC was associated with recurrence-free survival (RFS; Supplementary Fig. 9).

Dichotomization of samples based on sample-type specific median variant allele frequencies (VAFs), demonstrated an association between the level of ctDNA and NAC response for plasma collected both before and after NAC, but not for tumor DNA from urine supernatants and pellets (Supplementary Fig. 10). The level of ctDNA in plasma samples collected both before and after NAC and for utDNA in urine supernatants after NAC was furthermore associated with RFS (Supplementary Fig. 11).

### tdDNA liquid biopsy dynamics and treatment response

The tumor DNA dynamics during NAC (tdDNA either remained detectable or was cleared) for plasma samples remained significantly associated with NAC response(18) in this extended cohort (Fig. 4a). tdDNA dynamics for urine supernatants was also significantly associated with NAC response, but not for urine pellets (Fig. 4b-c). The response assessment for NAC reflects the local tumor response (pathological evaluation), so we next investigated the outcomes of the patients following RC to assess the response on distant micrometastatic disease.

**Fig 4.**
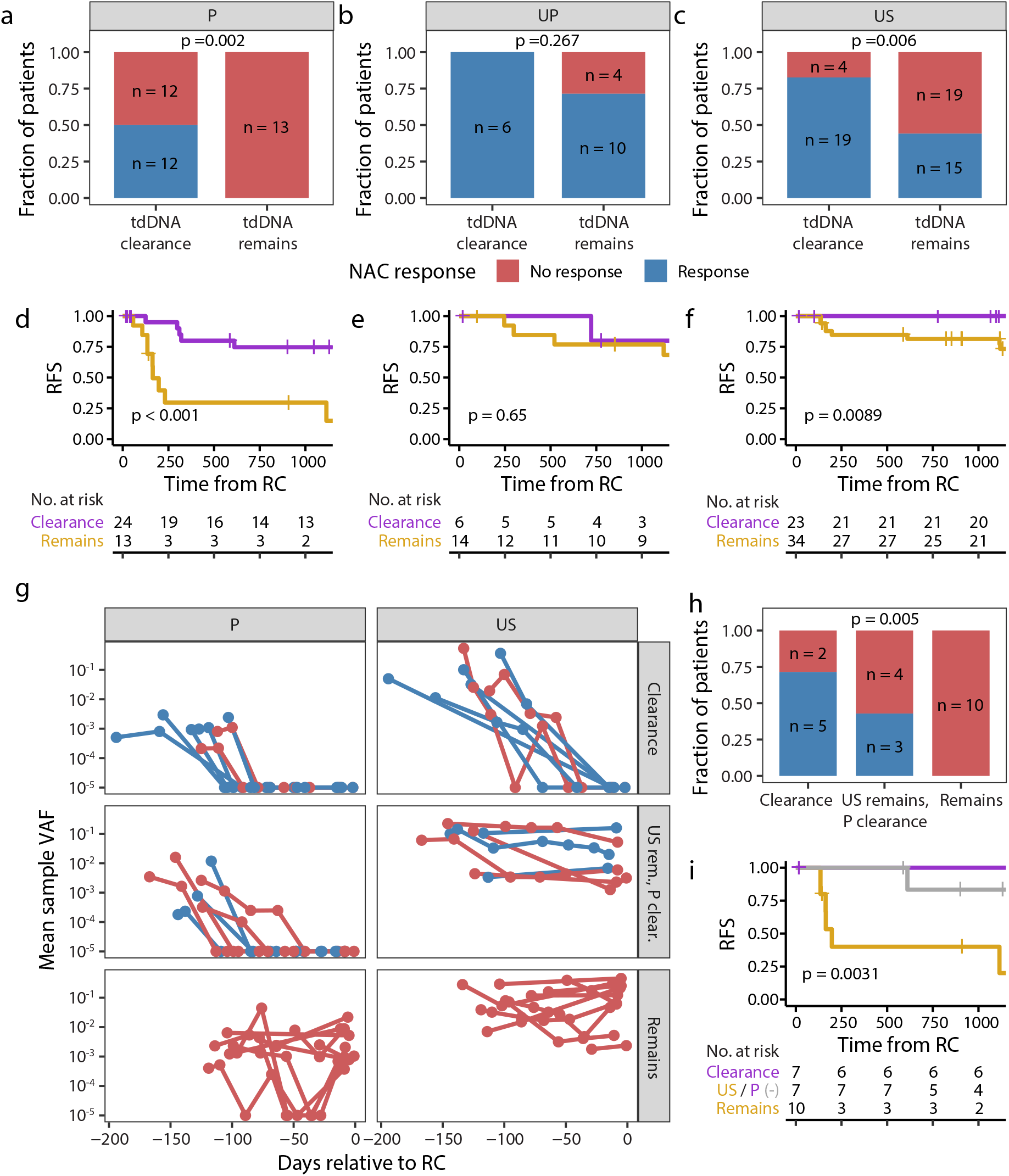
Tumor-derived DNA dynamics correlates with treatment response and outcome. a-c) Association between tumor-derived DNA (tdDNA) dynamics and neoadjuvant chemotherapy (NAC) response for a) plasma, b) urine pellet and c) urine supernatant. tdDNA clearance was defined as tdDNA going from detectable to non-detectable and tdDNA remains was defined as tdDNA remaining detectable. d-f) Kaplan-Meier survival analysis of tdDNA dynamics groups and recurrence-free survival (RFS) for d) plasma, e) urine pellet and f) urine supernatant. g) Detailed overview of tdDNA dynamics during NAC for patients with information available for both plasma and urine supernatant. Overviews were split according to sample type and agreement between sample-type tdDNA dynamics. h) Association between combined plasma and urine supernatant tdDNA dynamics and NAC response. i) Kaplan-Meier survival analysis of combined plasma and urine supernatant tdDNA dynamics groups and RFS.

Here we found that plasma tdDNA dynamics was strongly associated with RFS with particularly poor outcome for patients where tdDNA remained detectable (Fig. 4d). We did not observe an association between RFS and urine pellet tdDNA dynamics (Fig. 4e). For urine supernatants, as for plasma, we observed a strong association with RFS and a remarkable recurrence rate of 0% for patients with tdDNA clearance (Fig. 4f). Interestingly, a combination of tdDNA dynamics of plasma samples and urine supernatants showed concordance in 71% (17/24) of patients (Fig. 4g). The combination showed a potential for treatment response monitoring with concordant tdDNA dynamics being associated with response to treatment (Fig. 4h) and for risk stratification of patients, as demonstrated by the difference in RFS (Fig. 4i).

The overall effect of NAC was reflected by a decrease in detectable mutations in residual tumors from RC compared to the primary tumor. This was true for both WES and panel based mutation calls (Supplementary Fig. 12).

## Discussion

Treatment with NAC represents a clinical scenario in need of real-time monitoring to assess whether treatment is effective or initiation of alternative treatment would be optimal. Plasma samples have demonstrated promising utility based on a strong association between clearance of tdDNA and response to NAC, which importantly can be evaluated before RC is carried out(18). In addition, plasma tdDNA clearance has been associated with improved outcome following RC(18). These observations were further substantiated with the expanded patient cohort in this study.

Noteworthily, tracking of tdDNA in plasma samples during treatment is only possible in the subset of patients with detectable tdDNA before initiation of NAC treatment, which in this study amounted to approx. 43%. This number could also point to the patients that actually need systemic treatment because of disseminated disease. However, it is not clear how well the tdDNA detection methods actually capture micrometastatic disease. A recent study indicated that lesions have to be up to 10cm^3^ in size in order to be detected with current methods(26). The similar positive detection rates reached 85% and 89% for urine pellets and supernatants, respectively. This highlights that urine samples could provide a critical component in liquid biopsy based treatment monitoring for patients with MIBC treated with NAC simply based on the markedly higher detection rates. Importantly, we demonstrated an association between tdDNA dynamics in urine supernatants and a similar trend in urine pellets pointing to a potential future role for urine-based treatment response monitoring. Interestingly, we observed a particularly good outcome for patients with urine supernatant tdDNA clearance. This observation persisted when combining plasma and urine supernatant tdDNA measurements and highlights a potentially predictive and prognostic value of a combinatorial liquid biopsy based approach for monitoring of response, where a local- and systemic response measure is used in combination. However, the observations require validation in larger prospective cohorts.

Surprisingly, tdDNA was identified in both urine supernatants and pellets in samples obtained at visits with no concurrent tumors. This might be due to tumor lesions being missed or from release of tdDNA from mutant clones present in presumably normal appearing urothelium(27). However, recent studies using next generation sequencing or digital droplet PCR have identified tdDNA in urine supernatants obtained at visits with no concurrent tumor or concurrent low-grade non-invasive tumors(23,28,29). In addition, tdDNA was not identified in a subset of samples obtained at visits with MIBC. This has similarly been observed in a recent study by Chauhan *et al*.(19); however, the panel-based approach for tdDNA detection in this study could imply that the lack of tdDNA detection is for technical reasons, i.e. tumor-specific variants were not queried. Furthermore, urine samples represent a sample type with inherent variability in composition and concentration, which both could have implications for the level and detectability of tdDNA. Interestingly, we observed no variation in tdDNA detection rates and tDNA level across a wide range of cfDNA amounts. However, we did observe a correlation between cfDNA levels and several parameters assessed using urine dipstick data. The erythrocyte level was in line with this associated with the tdDNA call status of samples which aligns with haematuria as a frequent symptom of bladder cancer. Collectively, this demonstrates the extraordinary challenges created by the highly variable composition of urine samples. Future studies of urine-based liquid biopsy analysis should pursue further optimization of collection timing and procedures.

A limitation to the study is the different technologies applied for plasma tdDNA analysis. Both employ a patient-specific mutation selection approach based on WES data followed by targeted enrichment and sequencing, but differ in the enrichment strategies, variant calling and read-depth. Consequently, there are differences in the limit of detection of the applied methods. Importantly, all samples from single patients were analyzed with the same method.

In conclusion, we assess the potential for treatment response and outcome prediction by liquid biopsies collected in the pre-surgical setting for patients with MIBC. We validated previously observed prognostic and predictive features of plasma tdDNA analysis and similarly demonstrated prognostic and predictive power for urine supernatant tdDNA dynamics based on samples collected before, during and after NAC treatment. Importantly, tdDNA dynamics analysis provides a real-time assessment that could provide clinicians with information before RC is carried out.

Furthermore, plasma based tdDNA tracking is not feasible in a large fraction of patients in this setting, as tdDNA is either not being shed to the bloodstream or present at levels below the applied detection limit. However, urine-based samples provide the opportunity to track tdDNA in the vast majority of patients, and a combination of urine- and plasma based analysis may provide a robust measure of response as demonstrated in a subset of patients here.

## Data Availability

The raw sequencing data generated in this study are not publicly available as this compromise patient consent and ethics regulations in Denmark. Processed nonsensitive data are available upon reasonable request from the corresponding author.

## Acknowledgements

We would like to thank all technical personnel at the Departments of Molecular Medicine, Urology and Oncology, Aarhus University Hospital, for sample handling and processing.

