## Supplementary data for "Cell-free urine- and plasma DNA mutational analysis predicts neoadjuvant chemotherapy response and outcome in patients with muscle invasive bladder cancer"

### Whole exome sequencing

WES was performed using Twist Enzymatic Fragmentation Library prep and Human Core Exome Capture kit. Samples were sequenced on an Illumina Novaseq 6000. Samples from 56 patients included from Christensen et al.[1], were subjected to library preparation using Kapa HyperPrep and captured using SeqCap EZ MedExome or MedExomePlus. These samples were sequenced on an Illumina NextSeq 500. Raw sequencing data was initially processed using bcl2fastq2 and Trim Galore!/cutadapt (hg19/hg38 processing). FastQ files were processed according to the GATK Best Practices[2]: Alignment using bwa-mem, marking of duplicate reads using Picard, base recalibration using GATK, quality metrics were assessed using Picard. Mutations were identified using MuTect2 with default parameters except the threshold for maximum alternate alleles in the germline was raised. A custom filter selecting variants only vastly more present in the tumor and in regions with low noise, was subsequently applied[3]. Furthermore, variants identified by MuTect2 that did not pass the built-in filters were reintroduced if they were identified with high confidence using VarScan2/Strelka (hg19/hg38 processing)[4,5]. All somatic alterations were annotated using annovar[6]. Summarized metrics for WES data is available in Supplementary Table 2.

### Custom panel design

For selection, mutations were initially filtered to enrich statistically convincing mutations. A maximum of 1 mutation-containing read was allowed in the associated germline unless at least 20 times as many mutation-containing reads were identified in the tumor sample. Furthermore, a minimum read-depth of 10 for both the tumor and germline samples was set. We leveraged previously generated sequencing data using identical sequencing chemistry, but different genomic positions targeted (described in [7]). C>T/G>A mutations were frequently observed at positions with no expected mutations. In addition, the trinucleotide contexts "NCG" and "CGN" were frequently mutated. In samples from patients with MIBC, C>T mutations are the most commonly observed accounting for 51% of mutations in TCGA data[8]. Therefore, the top 10 mutations per patient based on the log odds score (TLOD) derived from

Mutect2[9], reflecting the probability of the variant being a true mutation, were prioritized regardless of the mutational context. Furthermore, up to 10 mutations assessed as damaging (Polyphen2: possibly/probably damaging or MutationAssessor: medium/high[10,11]), were included per patient in the panel. These were prioritized based on the variant allele frequency (VAF) and only included if the VAF was above the 1st quartile for the VAF of the sample in question in order to limit the selection of subclonal mutations. Similarly, up to 10 mutations in genes associated with bladder cancer (bladder cancer driver genes from [12] and significantly mutated genes in [8]) were selected if the VAF was above the 1st quartile. In addition, two TERT promoter mutations, which are frequently observed in patients with bladder cancer[13], were added to both panels. Collectively, the mutation selection process thereby serves to limit the error-rate of included genomic positions while optimizing the mutation selection to contain the mutations most likely to be clonal and impactful. Panel 1 was designed based on hg19 aligned WES data and panel 2 was designed based on hg38 aligned WES data. For comparative analyses, the panel 1 positions were carried over to hg38 using the R package rtracklayer[14].

**Custom panel variant calling pipeline**

The inclusion of genomic positions associated with multiple patients on every custom panel facilitates an abundance of sequencing data for every genomic position with no mutations expected to be present. This data can serve to build a background error model. To achieve this, we employed an analysis framework based on a maximum likelihood implementation of the shearwater algorithm developed by Gerstung et al[15]. Data generated from liquid biopsy samples was initially split by sample type before generating error models. For analysis of mutations for every patient, all data generated for a liquid biopsy type was loaded and other samples originating from the same patient were discarded. Following, the genomic positions associated with a patient were selected and base counts were generated for all samples of the given type. Only reads with a minimum mapping quality of 20 and bases with a minimum quality of 20 were considered. The presumably non-mutated data was fitted to a binomial distribution with site-specific calculation of the dispersion, as described by Gersung et al[15]. Presumably mutated data, i.e. data from the target patient, was assessed for a statistical significant difference compared to the background error model, resulting in p-values. Only base changes expected based on the mutation selection for the target enrichment were considered. Samples with a VAF above 25% were excluded when generating the error-model. Genomic positions with an average VAF above 10% across all considered samples and/or a read-depth below 50 in the target sample were excluded. P-values were corrected for multiple testing using the Benjamini-Hochberg procedure and adjusted p-values below 0.05 were considered significant.

The sample-wise tdDNA signal strength was assessed using Fisher's method on the target positions of a given patient and compared to the results of Fisher's method applied to a random selection of 50 mutations of all non-target mutations in the panel 10,000 times. A sample was categorized as tdDNA positive only if the results of Fisher's method was the relatively strongest when compared to the 10,000 random selections. Mean sample VAF was defined as the mean VAF of all mutations that were significant after adjusting for multiple testing. For samples with no significant mutations after adjusting for multiple testing, but with a sample-wise positive tdDNA call, the mean sample VAF was defined as the mean VAF for all mutations with unadjusted p-values below 0.1.

**Plasma-based ctDNA data using a 16-plex custom NGS approach**

Data for 288 plasma samples from 56 patients was obtained from a previous publication[1]. In brief, data was generated using WES data for tumor and germline samples and 16 presumably clonal mutations were selected for a custom NGS approach. Targeted sequencing was performed using an Illumina HiSeq 2500 to a read-depth of approx. 105,000x. Samples with at least 2 mutations identified, were considered tdDNA positive. Sample-level analytic sensitivity was previously determined to be greater than 95% at a 0.01% ctDNA level[16].

**Estimation of limit of detection**

We generated in-silico sample counts to assess the limit of detection (LOD) at every position for all included sample types. To accomplish this, we performed the initial steps of the shearwater-based calling algorithm[15], but replaced the counts from the target sample with in-silico counts representing 1-25 reads. A random sample was used to determine ratios between forward and reverse read depth for every position. The remainder of the calling pipeline was then carried out and the minimum number of reads required to reach statistical significance was determined for every position.

Based on the predetermined read depth of the in-silico sample, the LOD was then calculated (Supplementary Fig. 1). The specified LODs for the different sample types were inferred based on the mean LOD for the closest of the queried read depths. In addition, the LOD varied across the different base changes with C>T demonstrating the poorest LOD (Supplementary Fig. 1, read depth fixed at 3000X).

**Assessment of changes in the tumor mutational landscape during NAC**

We performed WES of DNA from tumor tissue obtained from RC specimens for 32 patients to assess the mutational changes that occurred during treatment with NAC. We observed a median of 708 mutations in these tumors and a median of 1295 mutations in the associated primary tumors. Surprisingly, only an average of 16.4% of mutations observed in primary tumors were detected in tumors from RC specimens (Supplementary Fig. 11). Similarly, a median of 19 (IQR: 0-31) of the panel mutations were identified in RC tumors (Supplementary Fig. 11). This was affected by a lower tumor cell percentage in RC tumors, as reflected by lower 90th percentile VAFs (Supplementary Fig. 11). Importantly, we found low 90th percentile VAFs for all RC tumors with none of the 50 mutations selected for targeted sequencing present. In addition, an average of 74% (range: 31-100%) of the observed mutations in RC tumors (selected for targeted sequencing) were present with VAFs above the 75th percentile of all mutations in the RC tumors pointing to enrichment of these mutations (Supplementary Fig. 11).

4. Koboldt DC, Zhang Q, Larson DE, et al. VarScan 2: somatic mutation and copy number alteration discovery in cancer by exome sequencing. *Genome Res*.

2012;22(3):568-576.

10. Adzhubei I, Jordan DM, Sunyaev SR. Predicting functional effect of human missense mutations using PolyPhen-2. *Curr Protoc Hum Genet*. 2013;Chapter 7:Unit7.20.

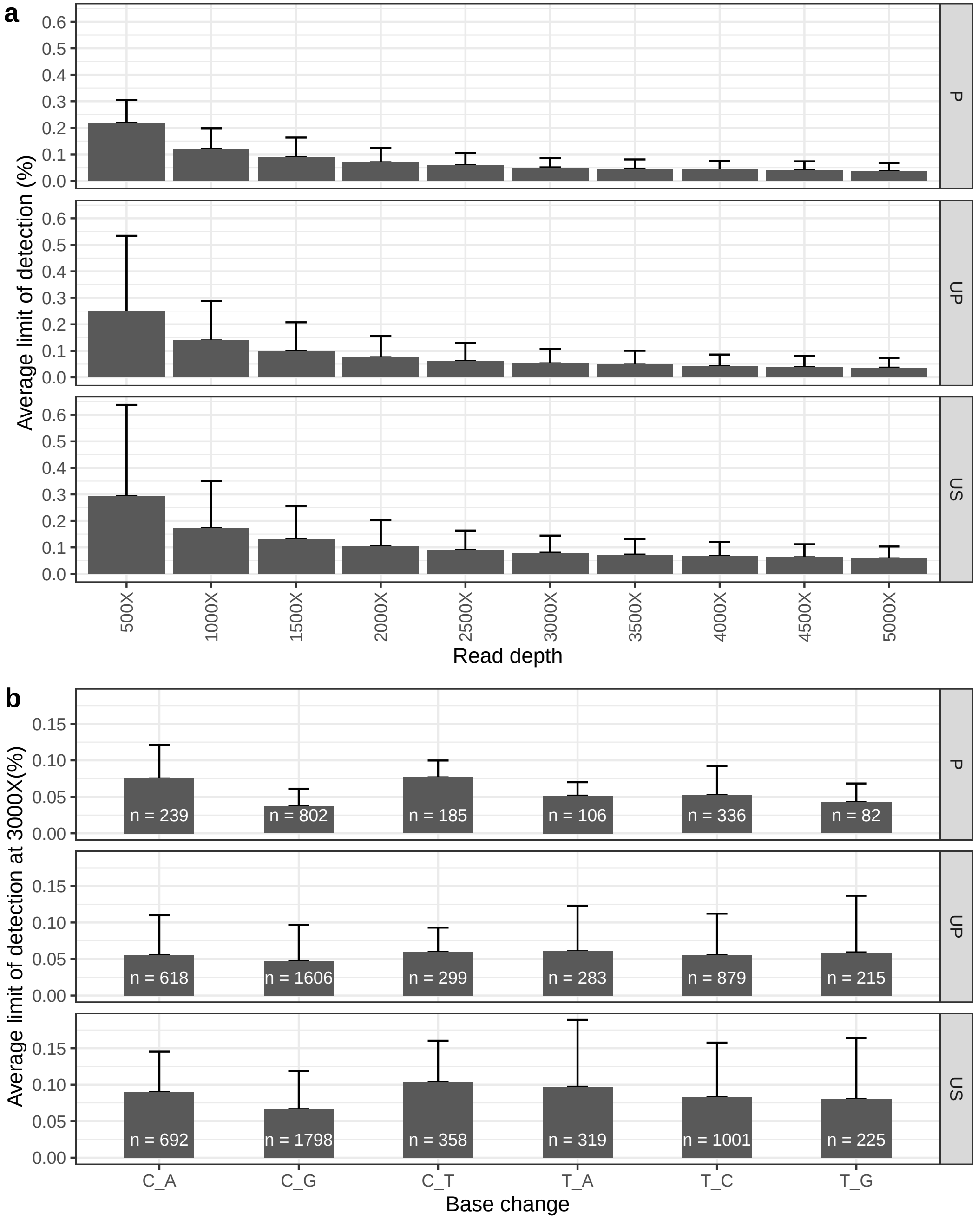


**Supplementary Fig. 1.**
In-silico analysis of the limit of detection for the applied methodology. a) Average limit of detection for all sample types by pre-specified read depths. Error-bars illustrate the standard deviation. b) Average limit of detection at a read depth of 3000X split by simple type and observed base change. Error-bars illustrate the standard deviation.

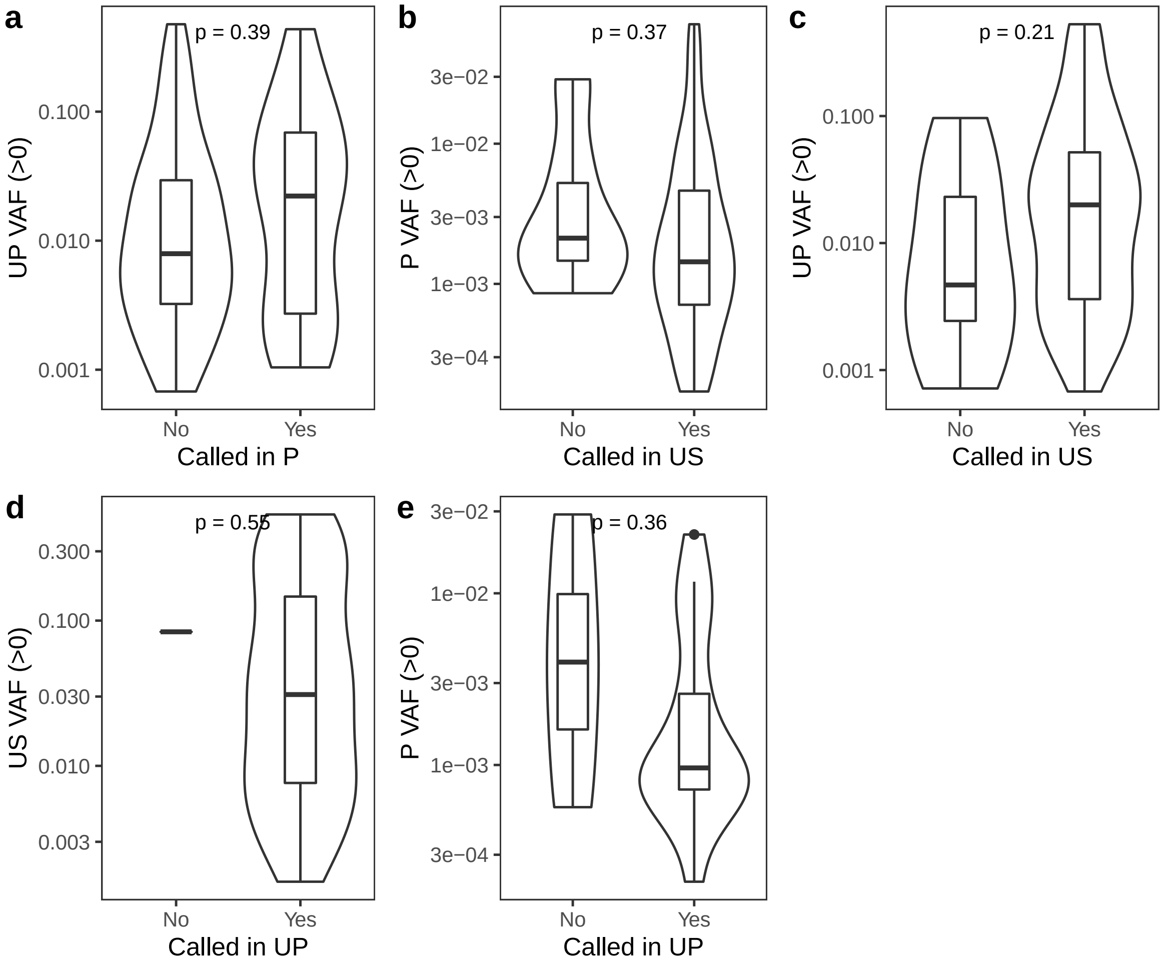


**Supplementary Fig. 2.**
Mean sample VAF levels for plasma, urine pellet and urine supernatant split by tdDNA call status in other paired sample types from the same clinical visit. a) Urine pellet level split by plasma call status. b) Plasma level split by urine supernatant call status. c) Urine pellet level split by urine supernatant call status. d) Urine supernatant level split by urine pellet call status. e) Plasma level split by urine pellet call status. Only samples with detectable tdDNA were considered. P-values were calculated using a wilcoxon rank sum test. P = Plasma, UP = Urine pellet, US = Urine supernatant.

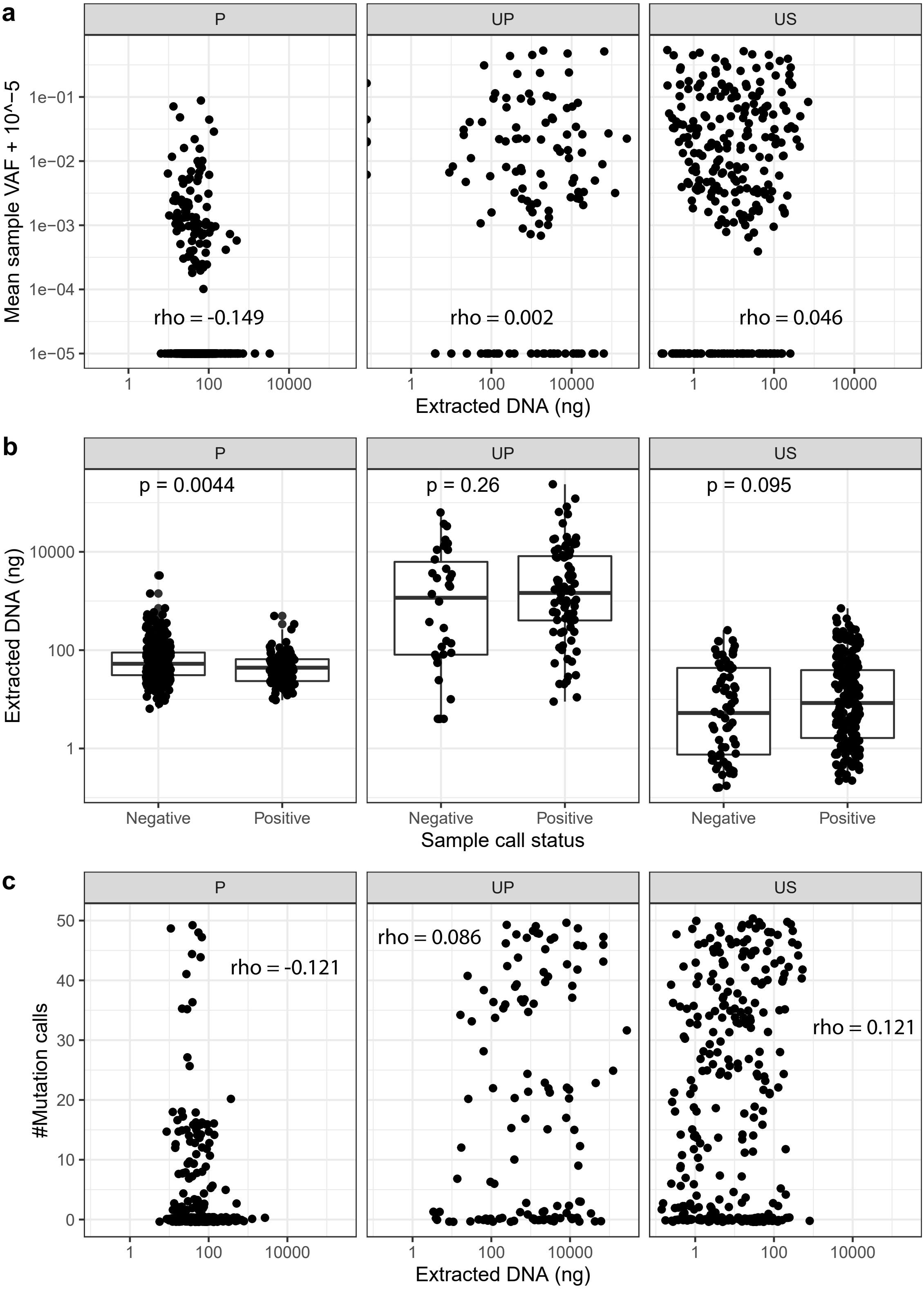


**Supplementary Fig. 3.**

Association between mean sample VAF (a), sample tdDNA call status (b) and number of tdDNA mutation calls (c) for plasma, urine pellet and urine supernatant and the amount of extracted DNA from the original sample. P-values were calculated using a Wilcoxon rank sum test and rho values were calculated using a Spearman correlation.

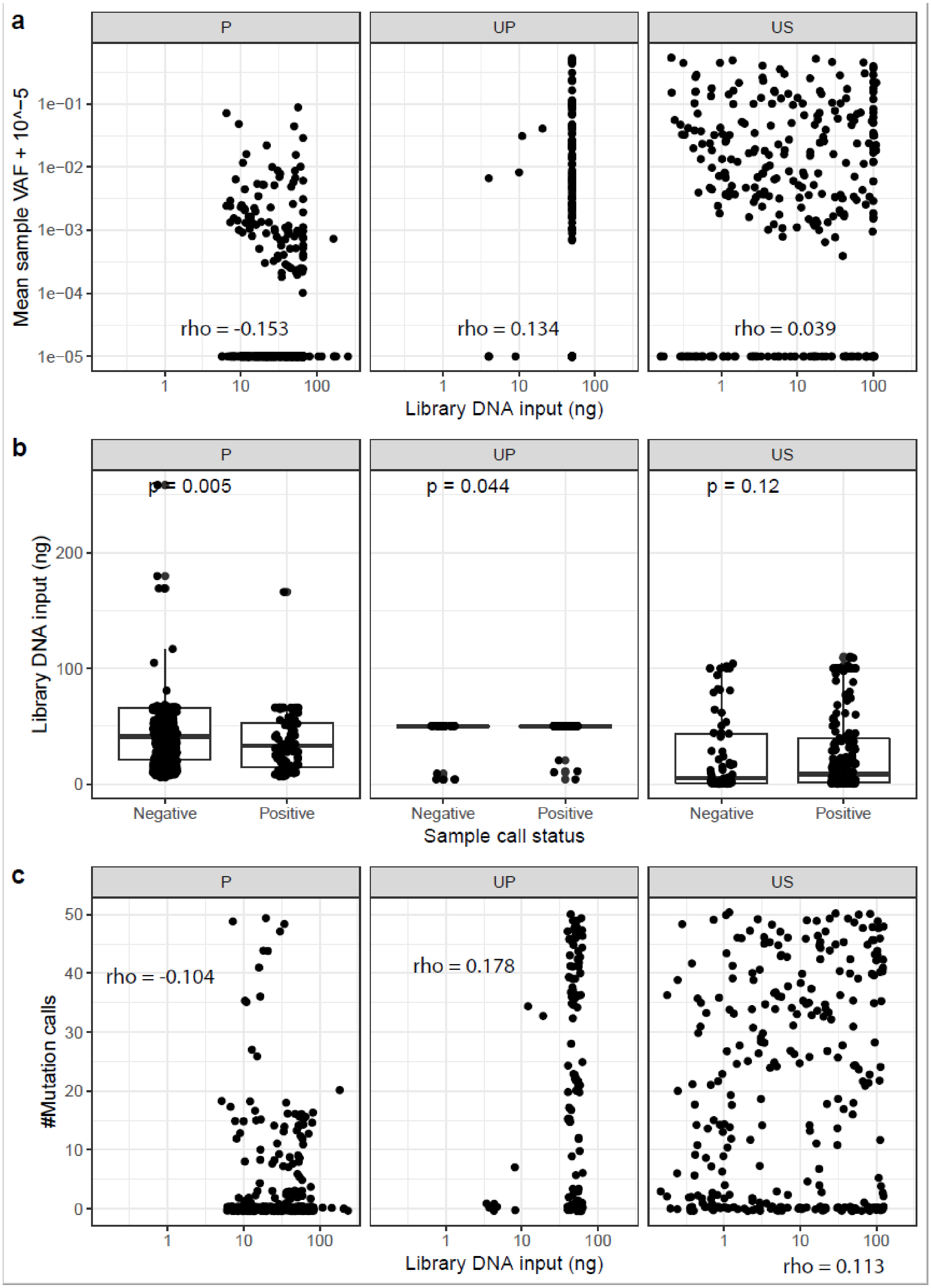


**Supplementary Fig. 4.**
Association between mean sample VAF (a), sample tdDNA call status (b) and number of tdDNA mutation calls (c) for plasma, urine pellet and urine supernatant and the amount of DNA used for NGS library input. P-values were calculated using a Wilcoxon rank sum test and rho values were calculated using a Spearman correlation.

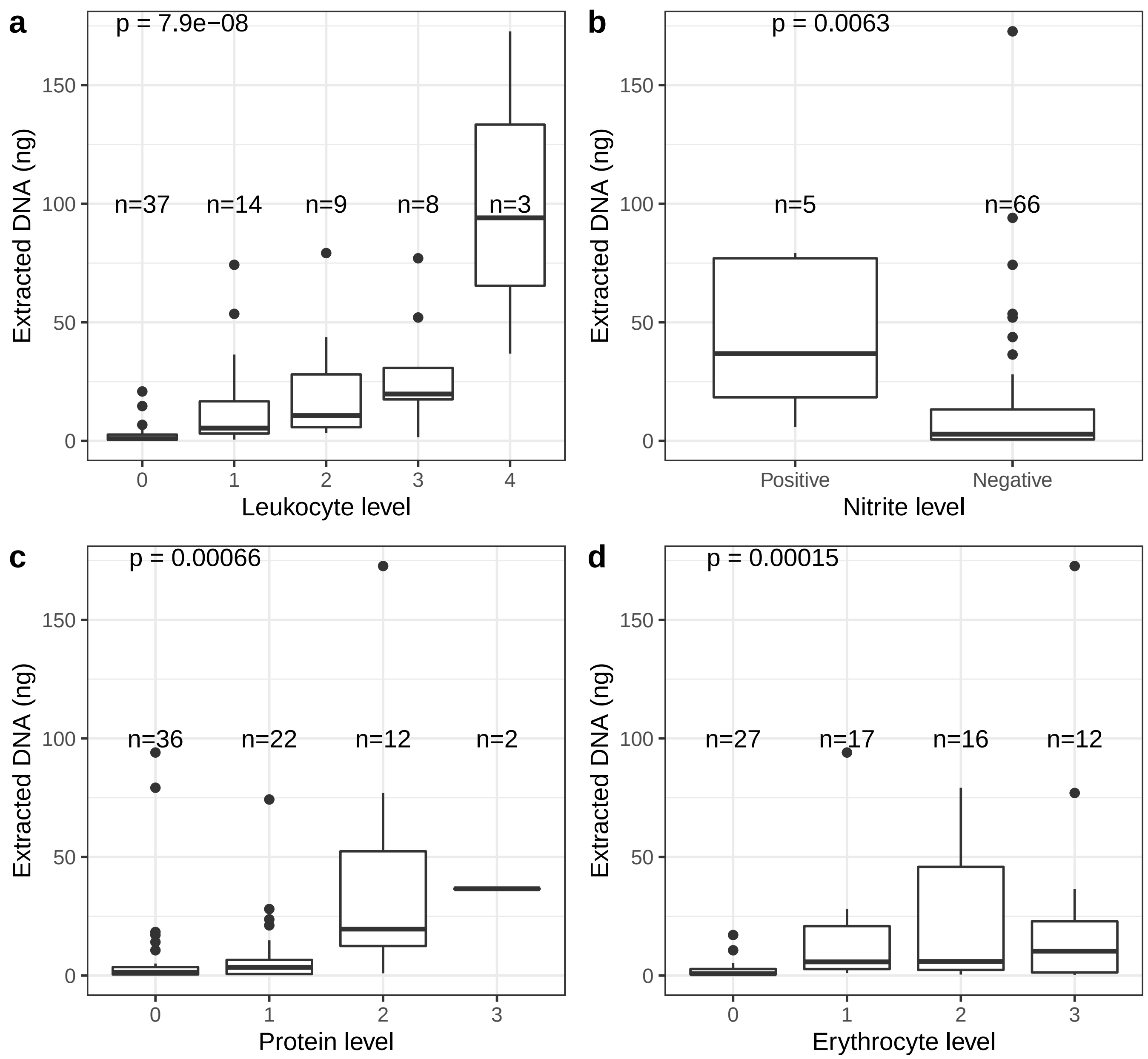


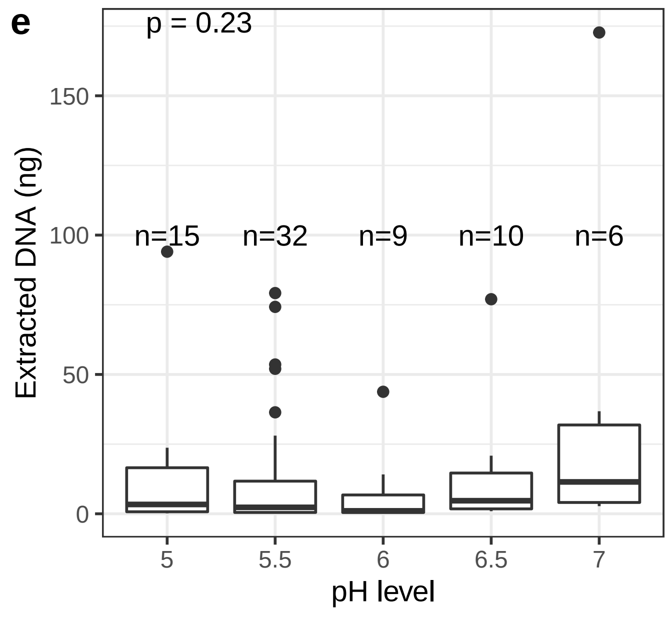


**Supplementary Fig. 5.**
Urine dipstick measurements compared to the amount of DNA extracted from urine supernatants collected at the same time (+/- 1 day). The following measurements were considered: Leukocyte level (a), nitrite level (b), protein level (c), erythrocyte level (d), pH level (e). P values were calculated using a Kruskal-Wallis test for comparisons with more than two groups and a Wilcoxon rank-sum test for comparisons with only two groups.

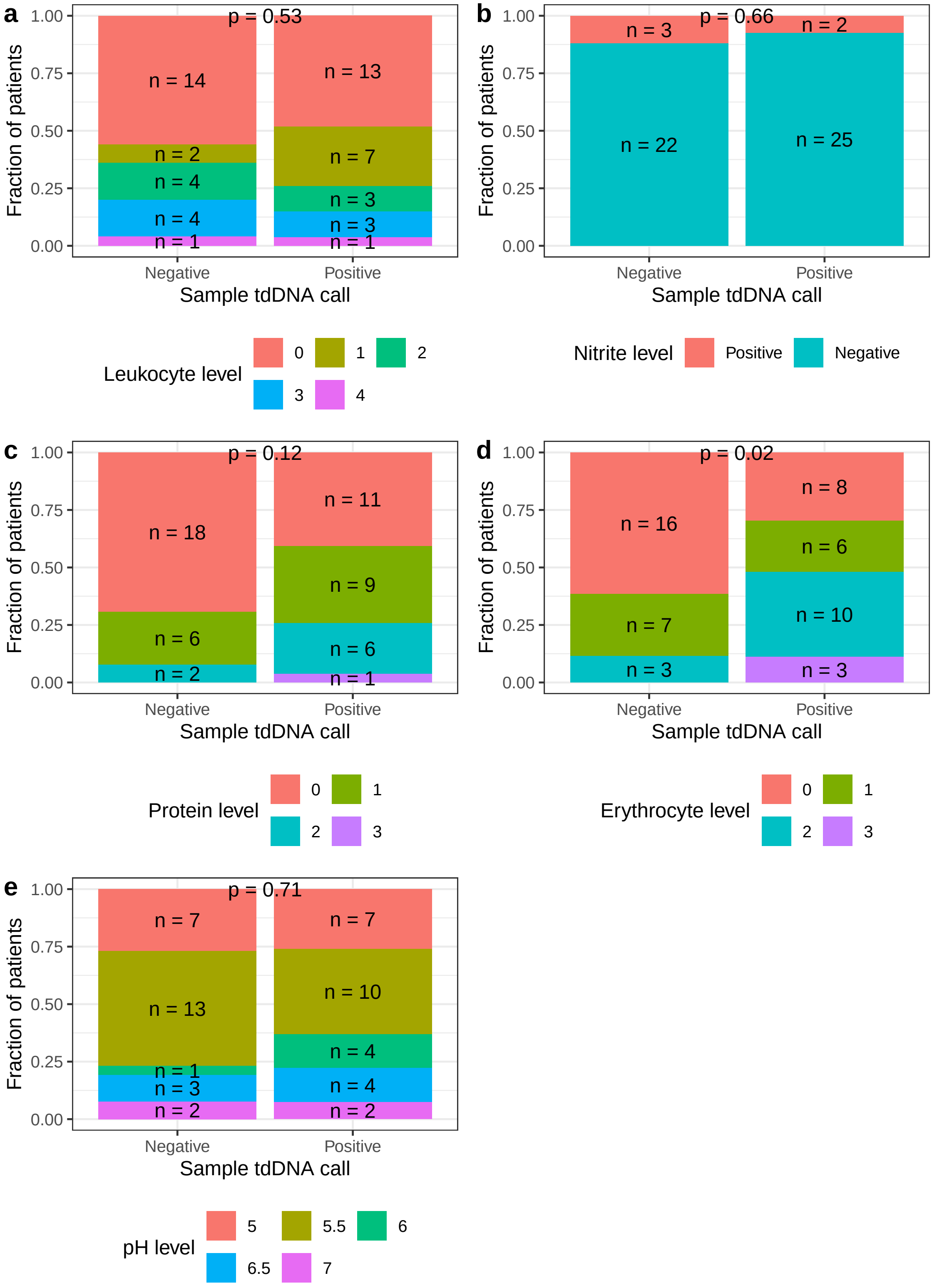


**Supplementary Fig. 6.**
Urine dipstick measurements compared to the tumor-derived DNA call status in urine supernatants collected at the same time (+/- 1 day). The following measurements were considered: Leukocyte level (a), nitrite level (b), protein level (c), erythrocyte level (d), pH level (e). P-values were calculated using Fisher’s Exact test.

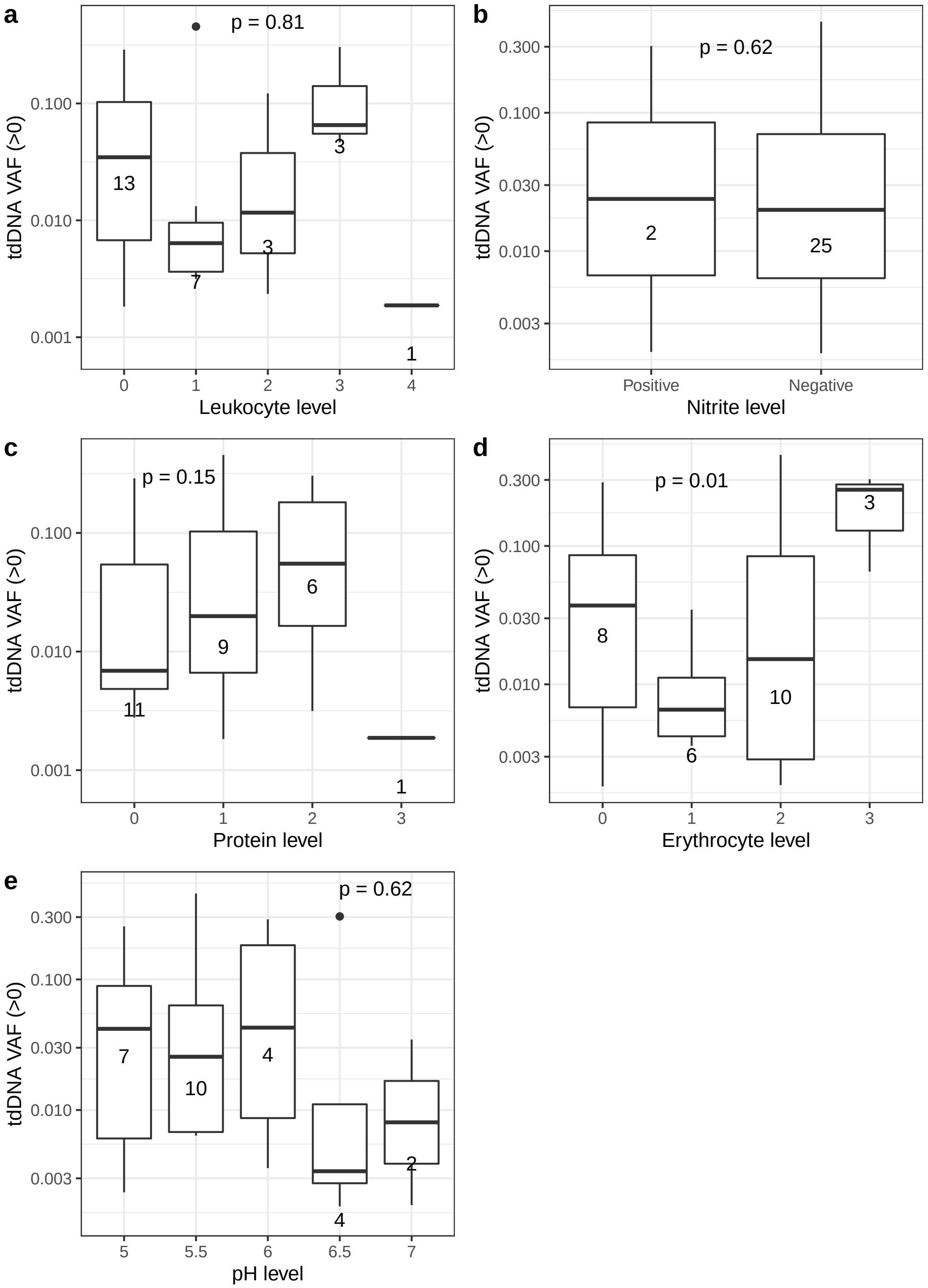


**Supplementary Fig. 7.**

Urine dipstick measurements compared to the tumor-derived DNA level in urine supernatants collected at the same time (+/- 1 day). The following measurements were considered: Leukocyte level (a), nitrite level (b), protein level (c), erythrocyte level (d), pH level (e). P-values were calculated using a Kruskal-Wallis test for comparisons with more than two groups and a Wilcoxon rank-sum test for comparisons with only two groups.

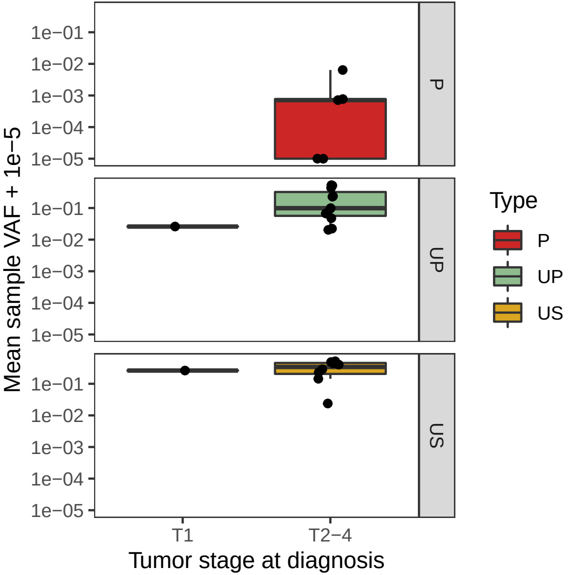


**Supplementary Fig. 8.**
Mean sample VAF levels for all patients with detectable tdDNA split by sample type. Tumor stage evaluation was based on the TURB specimen and tdDNA level based on samples collected before TURB.

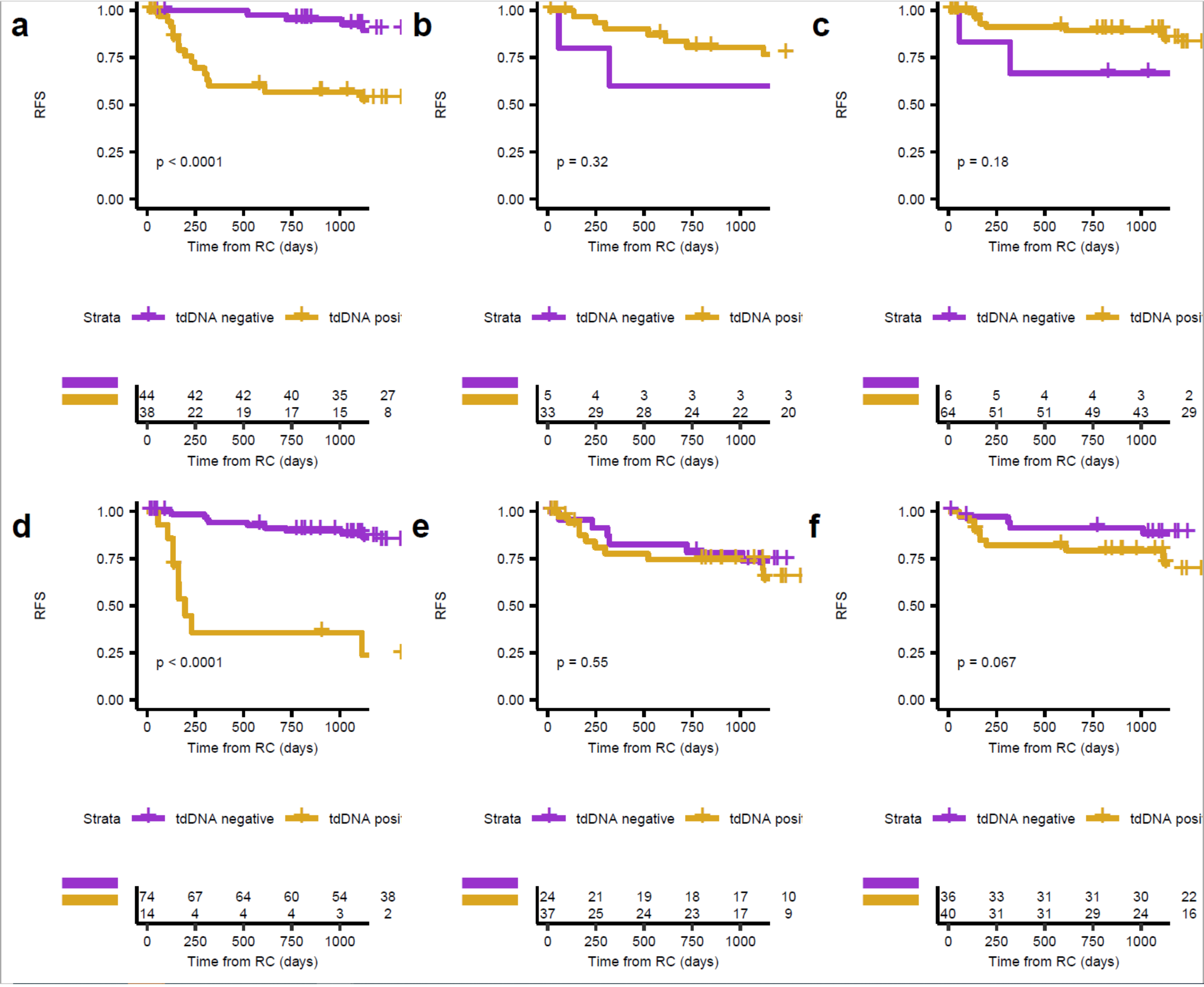


**Supplementary Fig. 9.**
Kaplan-Meier survival analysis of recurrence-free survival stratified by tdDNA call status for all sample types. a-c) tdDNA call based on samples collected before treatment with NAC for a) plasma, b) urine pellet and c) urine supernatant. d-e) tdDNA call based on samples collected after treatment with NAC for d) plasma, e) urine pellet and f) urine supernatant.

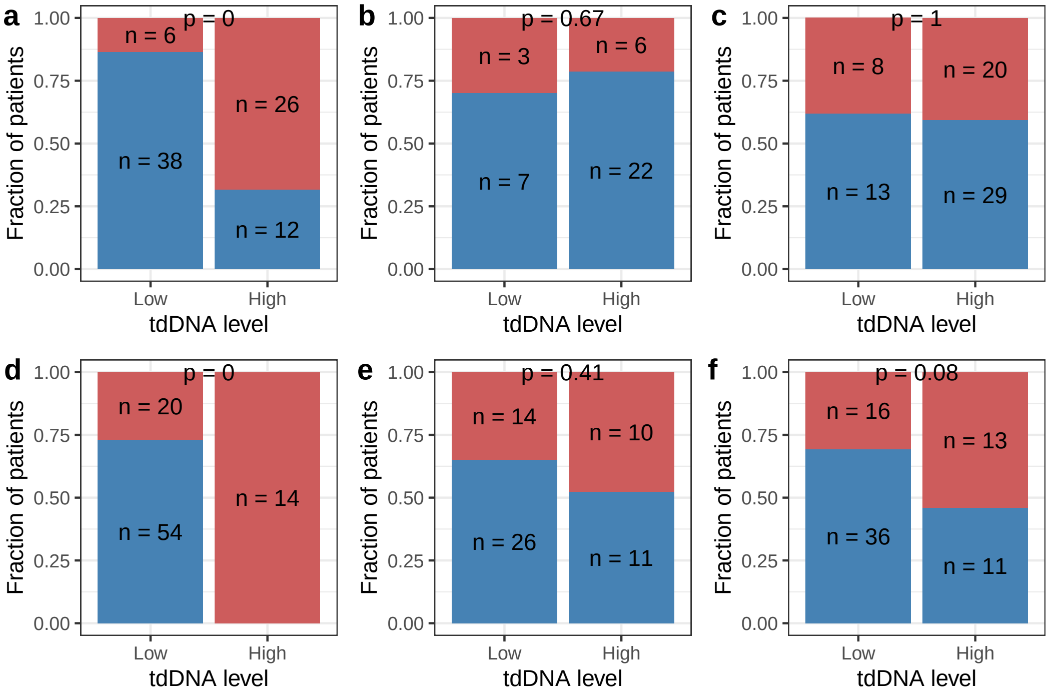


**Supplementary Fig. 10.**

tdDNA level compared to NAC response. a-c) tdDNA status for samples collected before NAC compared to NAC response for a) plasma, b) urine pellet and c) urine supernatant. d-f) tdDNA status for samples collected after NAC compared to NAC response for d) plasma, e) urine pellet and f) urine supernatant. P-values were calculated using Fisher's Exact test. Red indicates no response and blue indicates response to NAC treatment. tdDNA level was determined by dichotomization based on sample-type specific median values.

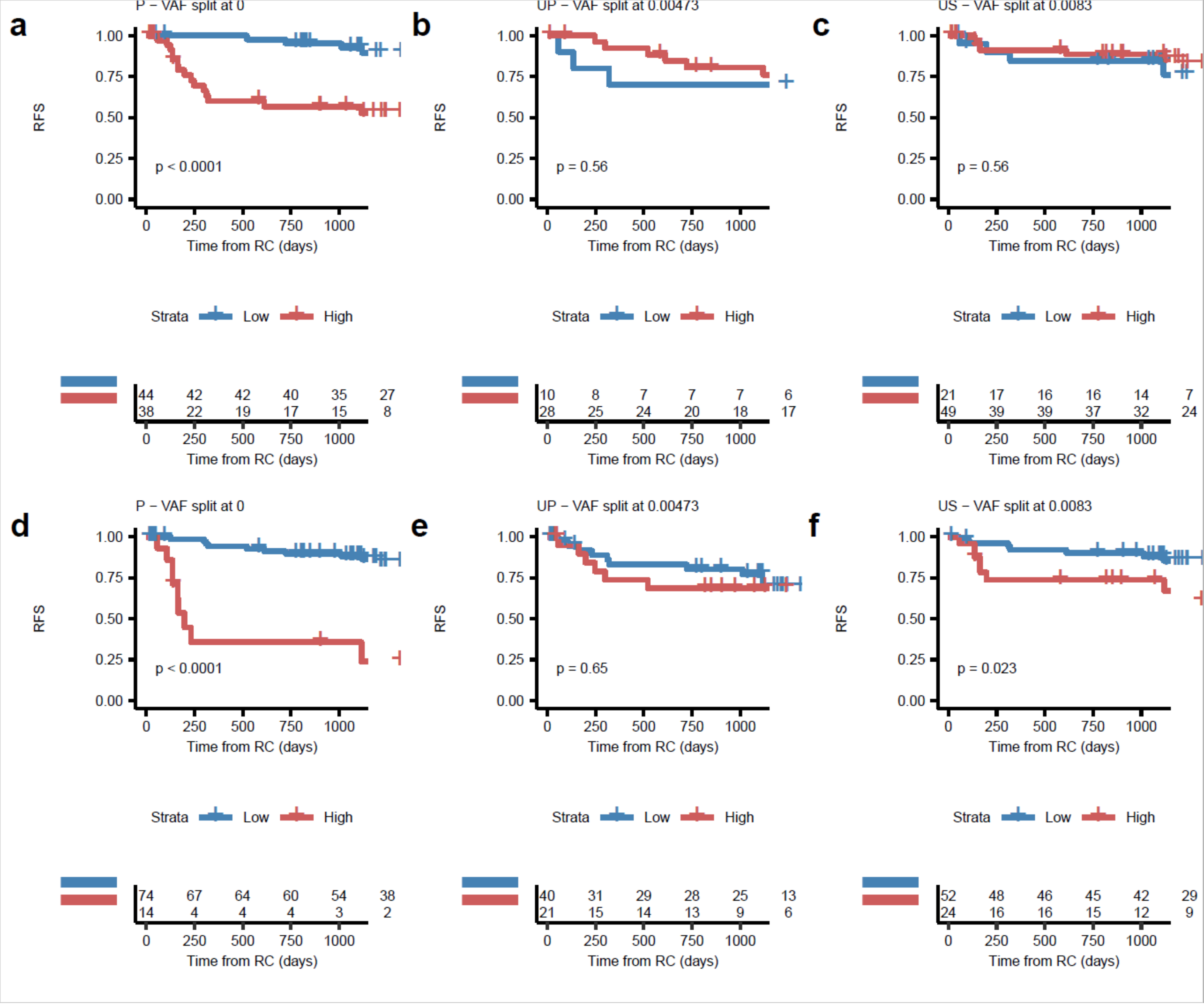


**Supplementary Fig. 11.**
Kaplan-Meier survival analysis of recurrence-free survival stratified by tdDNA level for all sample types. a-c) tdDNA call based on samples collected before treatment with NAC for a) plasma, b) urine pellet and c) urine supernatant. d-e) tdDNA call based on samples collected after treatment with NAC for d) plasma, e) urine pellet and f) urine supernatant. tdDNA level was determined by dichotomization based on sample-type specific median values.

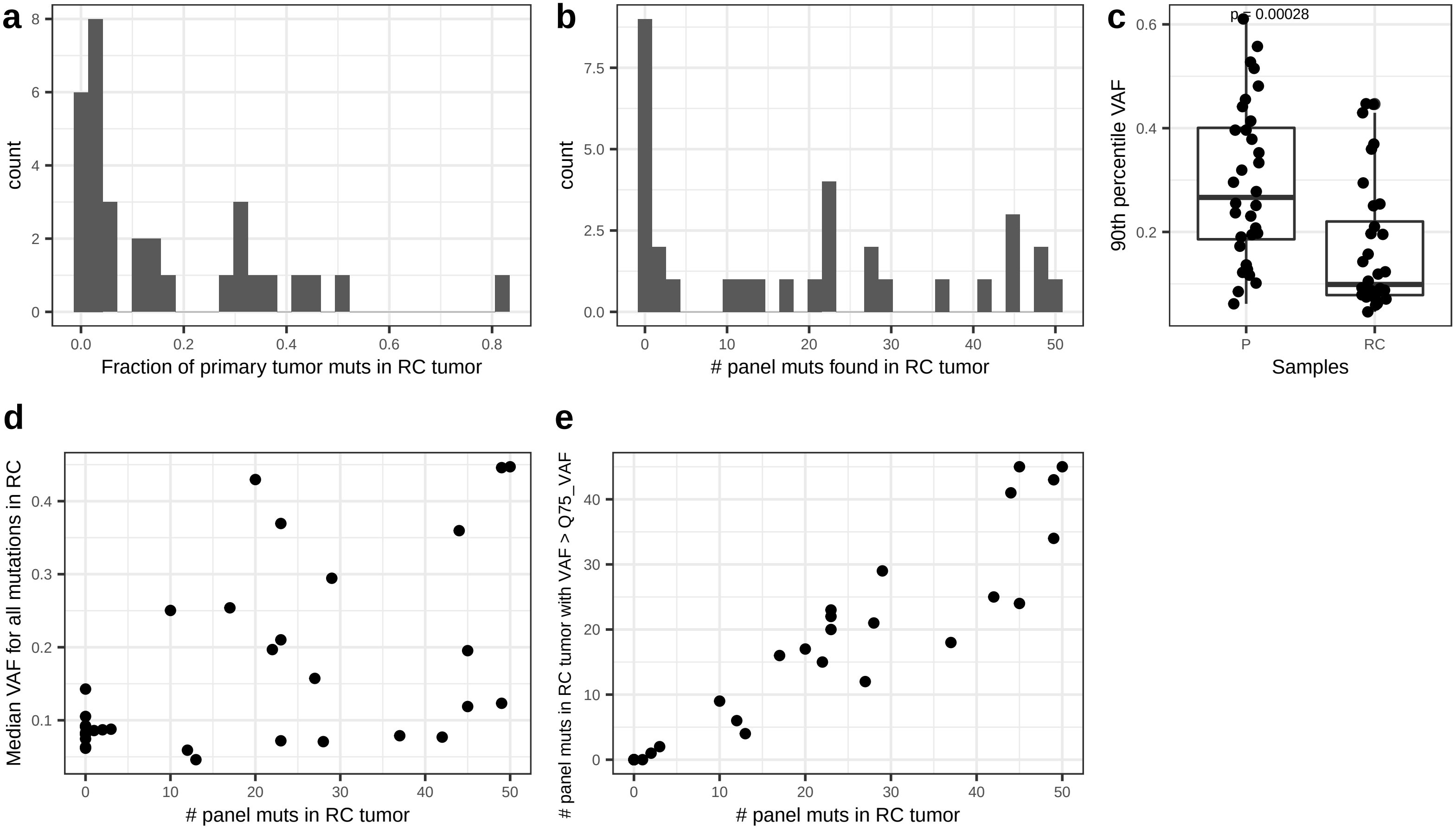


**Supplementary Fig. 12.**

Comparison between mutations observed in primary and residual tumors at RC. a) The fraction of mutations observed in the primary tumor that was also detected in the associated residual tumor from WES. b) Number of custom panel mutations detected per patient in the paired residual tumor. c) The observed 90th percentile VAF levels for primary and residual tumors. Only paired tumors are illustrated and the p-value was obtained using a Wilcoxon Signed-rank test. d) Association between the median VAF for all mutations detected in residual tumors and the number of custom panel mutations detected. e) Association between the number of custom panel mutations with a VAF above the 75th percentile VAF and the number of custom panel mutations detected in residual tumors.
